# Evaluation of Self-Directed Learning Activities at King Abdulaziz University: A Qualitative Study of Faculty Perceptions

**DOI:** 10.1101/2024.10.20.24315769

**Authors:** Ammar Balkheyour, Michal Tombs

## Abstract

**Purpose:** Doctors are expected to be lifelong learners and engage in continuous professional development throughout their careers. Learning to be a self-directed learner as early as possible is therefore likely to lay the foundation for future learning and development. For this reason, Self-Directed Learning (SDL) has recently been incorporated into the internal medicine curriculum at the Faculty of Medicine in Rabigh at King Abdulaziz University, Saudi Arabia. The purpose of this study was to evaluate faculty perceptions of the effectiveness of these SDL activities.

**Methods:** The evaluation framework for this study was underpinned by Stufflebeam’s Context, Input, Process and Product (CIPP) evaluation model. Seven faculty members took part in semi-structured interviews that explored their understandings of SDL objectives (Context), their perceptions of the methods and resources used (Input), the implementation of SDL activities (Process) and whether they meet their intended educational goals (Product). Interviews were transcribed and analysed using the thematic analysis method.

**Results:** Four themes emerged from data and were mapped against the elements of CIPP model: these themes are as follows: faculty perception of SDL (context), content and resources (input), facilitation and scheduling (process) and student engagement and assessment (product).

**Conclusions:** The faculty had different opinions about the objectives and content of SDL sessions; however, they agreed that these are aligned with internal medicine objectives and clinical content. Faculty perceived SDL activities as a reading task for students to complete prior to group discussion. The data revealed the necessity for faculty training to conduct such sessions. Challenges in the learning environment were reported, including issues in the library access and scheduling of the academic activities. Participants reported poor engagement from students to be a particular challenge and have made suggestions on how this could be addressed. In addition, they emphasized the need for assessment for these sessions.

## Introduction

The Faculty of Medicine in Rabigh, King Abdulaziz University in the Kingdom of Saudi Arabia has undergone a significant shift in the educational system moving from a teacher-centred to a learner-centred model (1). The internal medicine curriculum has subsequently changed to reflect this shift, moving as much as possible from traditional lectures and bedside teaching to a more hybrid model, with the addition of activities, case-based learning (CBL), evidence-based medicine and clinical reasoning skills with an emphasis on self-directed learning (SDL). Medical doctors are required to continuously learn in a field where knowledge is being rapidly updated, and for this reason, SDL is perceived to be for lifelong learning and professional development (2, 3). Knowles (4) defined SDL as ’a process in which individuals take the initiative, with or without the help of others, in diagnosing their learning needs, formulating goals, identifying human and material resources for learning, choosing, and implementing appropriate learning strategies, and evaluating learning outcomes’.

SDL is an active research area and research suggests that enabling students to develop self-directed learning skills has the potential to enhance academic achievement and career development (5). However, findings highlight that the success of such activities and the effectiveness of their implementations greatly depends on students’ and teachers’ attitudes and perceptions of SDL (6).

Students’ and faculty’s perceptions of SDL have been studied worldwide, with most studies examining whether students have the necessary self-directed skills and exploring factors associated with SDL (7, 8). Such research typically asks students to self-report their level of SDL skills using a measure such as Guglielmino’s SDLR (self-directed readiness) scale and Fisher et al.’s SDLR scale.

Research suggests that students report higher SDL skills in the later years of their studies, and that certain personality traits such as independence, willingness to learn, and individual learning accountability are positively linked with scores on the SDLR (7–9). In Saudi Arabia, research conducted with medical students suggest that they are motivated, but they are deficient in time management skills, and they require additional support and guidance from tutors to become more self-directed (5, 10, 11). However, few studies evaluated SDL interventions or examined the implementation of SDL-based activities or programmes.

One of these evaluative studies, a study was conducted at Saint Boniface School of Nursing in Manitoba, Canada (12). The research used Guglielmino’s SDLR scale and the Conti Principles of Adult Learning Scale to evaluate students’ readiness and faculty’s use of adult learning principles. Results showed a positive relationship between the student age, formal education, and SDLR scale scores. The study recommended focusing on developing SDL skills for students, and incorporating problem-solving in nursing education. The study also highlighted the importance of educators as facilitators of learning.

More recently, a study by Hill, Peters (13) evaluated the effectiveness of a case-based SDL activity in a pre-clinical course for first-year medical students.

The study used the TBL (team-based learning) approach, delivering case studies in microbiology over a 6-week course. The researchers concluded that the implementation of case-based SDL in the form of TBL was valuable for first-year students. Another study in Denmark evaluated self-directed CECs (clinical encounter cards) as a self-directed learning tool for undergraduate medical students (14). Results showed positive effects on engagement in diagnostic reasoning, reflection on management plans, and professional identity formation. However, students rated CECs as having low usefulness with preceptor support, with most comments being negative. This study suggests that CECs can be effective in certain contexts and highlights the facilitator role in SDL.

Locally, perceptions of students were examined on the main campus of King Abdulaziz University (15) and its branch at the Faculty of Medicine in Rabigh (5). The results of both studies aligned with findings from other Saudi institutions.

The Faculty of Medicine at King Abdulaziz University’s main campus assessed students’ SDLR using Guglielmino’s SDLR scale. The study found that over 60% of respondents believed SDL would improve learning, with the most common feature being a lack of resources and personal ability. However, 99% of respondents were below average in readiness for SDL. In the other study, a qualitative focus group study at the Faculty of Medicine in Rabigh evaluated the student experience with SDL and its effectiveness for lifelong careers. The study included 29 students from different academic years and focused on five themes: understanding SDL, views of SDL as a strategy, process of the strategy, effects of SDL, and SDL and lifelong learning. Most students enjoyed SDL, but time management was a challenge.

There are regular, ongoing evaluations of students’ satisfaction at the Faculty of Medicine in Rabigh. However, no formal evaluation has been conducted for the newly implemented SDL activities. In addition, internal medicine modules at the Faculty of Medicine in Rabigh have explicitly implemented SDL in their curriculum, which is unique from other educational strategies reported in previous studies such as PBL (problem-based learning), TBL and CECs (13–16). Moreover, the literature lacks studies investigating SDL from a faculty perspective in Saudi Arabia, although previous research has claimed that faculty facilitation has an essential role in the SDL process (12, 17, 18) . Additionally, the success of an evaluative study largely depends on the participation of stakeholders (19) and faculty are primary stakeholders; hence, their opinions are needed for further development and improvement. Therefore, there is a need to investigate the unique experience of the Department of Internal Medicine from the faculty perspective considering the context and multiple factors affecting SDL Several evaluation models exist to assist evaluators in choosing the appropriate methods for their specific evaluative questions (20). One of the most cited models is the Context, Input, Process and Product (CIPP) model by Stufflebeam (1971). This evaluative study was based on the CIPP model of evaluation. The research aims to evaluate the effectiveness of SDL activities currently delivered to internal medicine students at the Faculty of Medicine in Rabigh, King Abdulaziz University, Saudi Arabia using the CIPP evaluation model from the perspective of the faculty who are involved in delivering SDL sessions.

Therefore, the study objectives were to examine faculty perceptions regarding the following:

1. What are faculty’s understanding of SDL activities and their objectives? (Context)
2. What are faculty perceptions of the steps and resources used (Input), and what are the potential challenges in their implementation? (Input and Process)
3. Do SDL activities meet their intended educational goals and do they address students’ needs and improve their ability to be self-directed? (Product)

## Methods

### Study design

A qualitative methodology, through semi-structured interviews, was used as the aim of the study was to inform the local context and not to generalise findings. Its focus was on exploring faculty’s personal experiences within the context of the school, seeking to generate rich data and a deep understanding of the phenomena being studied (22).

### Sample

Seventeen faculty members at the Department of Internal Medicine, Faculty of Medicine in Rabigh are actively involved in delivering SDL sessions. All these members were invited to participate in the study. An invitation letter was sent to the distribution e-mail list of the Department of Internal Medicine, Faculty of Medicine in Rabigh and through a social network (the WhatsApp group for the module). 7 faculty members volunteered to take part in the interviews, including 5 males and 2 females aged 36–65 years. Participants were assistant professors and professors. Their experience in teaching ranged from 2 to 20 years, and their experience in delivering SDL in the target department ranged from 1 to 5 years.

The interviews were mainly face-to-face; however, two interviews were conducted via videoconferencing (online). The interviews were 35–48 minutes in length.

### Materials and Procedure

An interview guide was developed to include questions that were mapped against the CIPP model of evaluation (21). Questions were grouped into two sections. The first section was aligned to the Context and Input components, asking about their understanding of SDL activities and their objectives as well as their educational strategies when delivering these sessions. The second section focused on the Process and Product components, and included questions related to SDL implementation challenges and whether they meet their educational goals. The interview guide was revised after the first interview. The personal data collected in this study were age, sex, academic rank, years of academic experience, and number of years of delivering SDL sessions.

Participants were given a participant information sheet and consent form to read and sign prior to taking part in the interview. Five face-to-face interviews and two online interviews were conducted, all were one-to-one and about 40 minutes in duration. The interview guide with a list of questions and their sequence in addition to more revealing follow-up or probing questions to obtain further data, were used; however, the order of the questions was responsive to the participants’ interactions (23, 24). All the interviews were audio recorded and transcribed to analyse the conversations.

### Ethics approval

The study was ethically approved by the School of Medicine Research Ethics Committee at Cardiff University (SMREC reference number 22/59) and permission from the gatekeeper, who is the chairman of the Department of Internal Medicine at the Faculty of Medicine in Rabigh, King Abdulaziz University was obtained. Informed consent was taken from all participants and signed by them before the start of each interview.

## Results

Data were analysed using the thematic analysis method (25) and presented as four themes (Table 1). Each theme represents an element of the CIPP model. These themes are as follows: faculty perception of SDL (context), content and resources (input), facilitation and scheduling (process) and student engagement and assessment (product).

**Table 1:** Thematic analysis results.

|  | **Theme** | **Description** |
| --- | --- | --- |
| 1 | **Context evaluation:**<br>**Faculty's perceptions**<br>**of SDL** | Faculty's understanding of SDL and perceptions of their objectives and preparedness for SDL activities |
| 2 | **Input evaluation:**<br>**Content and resources** | Faculty perceptions of topics, content and resources used in SDL sessions |
| 3 | **Process evaluation:**<br>**Facilitation and**<br>**Scheduling** | Perceptions of the facilitation process and scheduling |
| 4 | **Product Evaluation:**<br>**Student engagement**<br>**and assessment** | Perceptions of the sessions in achieving its intended outcomes and suggestions for improvements |

### Theme 1: Context evaluation - faculty’s perceptions of SDL activities and their objectives

Context evaluation considers the need, problem, and opportunity within a situation (26). To this end, responses were analysed to examine whether faculty had a clear understanding of the objectives and purpose of SDL sessions and whether they felt prepared to facilitate them.

Participants’ responses indicated that most perceived it to be a two-step activity, including a reading task to be completed by the students in preparation for the second facilitation part. For example, one participant stated ‘*[SDL] is a task you give students, such as an article, summary, or Internet link to read. Then I meet with them either online or in the class to go through discussion, and I ask questions to check their understanding’.* Another participant also stated that ‘*self-directed learning is that the students read the topic and understand it by themselves, then we’ll present the topic to their colleagues and to the instructor, and then there will be a discussion between the students and the instructors about this topic’.* This was also echoed by a third participant who described it as follows: *’First, I choose a certain disease and related updated guidelines. Then, I send the article to the students one week before. After that, they are requested to summarise the important points in the management guidelines’*.

When asked about the objectives of these activities, all participants agreed that the SDL sessions were aligned with the internal medicine module objectives in terms of the clinical topic. However, they were not aware as to whether the curriculum for internal medicine contained objectives specific to SDL skills. One participant mentioned: *’I am not aware of any SDL objectives. There is a blueprint that includes objectives for the whole module… ’*. One participant did recognise that *’the main target is to improve the confidence in students to make them more independent’, whilst others* stated that the objective of SDL sessions *‘teaching the students how to utilise online resources and read articles*’ Almost all participants believed that SDL sessions addressed the students’ need to be more self-directed in their studies and noticed improvement in the students’ ability to acquire knowledge in other sessions, such as evidence-based medicine and bedside teaching sessions. For example, one participant stated: *‘Yes, somehow, I noticed in evidence-based medicine sessions that they knew how to search the literature, randomised control trials and systemic reviews. They learned how to read these articles. I noticed some improvement in the sixth year’* All participants mentioned that they did not receive any formal instructions regarding SDL sessions and some reported that they did not know how they should run them. One participant mentioned, ’*I don’t know how ideally SDL should be taught’, and another* participant stated, *’No, I was not given any formal instructions or guidelines. I was told some instructions verbally, but I did not understand them clearly’*. Faculty training was frequently recommended by the interviewees, suggesting that this can be achieved by arranging a course or workshop that helps instructors be better prepared as noted by one participant: ’*We need workshops from the Medical Education Unit to help us improve our delivery of the modules, whether for SDL or other types of sessions’*. One of the participants suggested peer review as a method for faculty training for SDL, stating, ’*faculty can be monitored by other faculty who act as trainers while delivering SDL sessions’*

### Theme 2: Input: Content and resources

Input evaluation assists in the decision-making of how facilities and resources will be determined in order to achieve the goal of education (26). This theme therefore considered faculty perceptions regarding the content and the learning environment.

In particular, the theme demonstrates how faculty select the topics and content for the SDL session and their perceptions of the resources and the learning environment.

Some of the responses indicated that faculty prefer to choose topics that are not covered in other teaching activities (e.g. lectures) but are aligned to the module objectives and relevant clinical practice, as stated by one participant, *’I choose a topic not covered in the lectures that are clinically relevant. I like to focus on disease management’*. Other participants on the other hand preferred selecting their content based on the knowledge and topic area taught in the lectures in four different ways. First, the SDL session could describe practical guidelines for one of the diseases covered in the lectures. For example, one participant mentioned, *‘I choose an article that is landmark and practice changing that helped in updating guidelines…I prefer it to be about one of the diseases covered in the lectures’.* Second, the tutor may include references to read for some of the knowledge informed in the lectures. Third, the tutor utilises SDL sessions to cover points planned to be covered in the lectures that were omitted due to time constraints, as reported by one participant: ***‘****Usually, I choose the content that I didn’t have time to cover during the lectures. For example… We talked about the risk factors briefly, but the level of evidence for each risk factor was not covered because it’s a long subject’.* Fourth, SDL sessions are based on one of the topics that is considered important or on a common health problem as mentioned by one participant: *‘I decide according to the importance of the subject. For example… I pick up a common disease… so it depends on the importance and how common the disease is for that week’* Generally, participants felt satisfied with the available technology and online platforms for their SDL sessions. One participant did not prefer online sessions because it does not allow for direct interaction with the students: *’Yes, I don’t like online sessions. Many students don’t turn on the camera, so we don’t see if the student is following the session or not. Sometimes, the Internet is interrupted. You don’t know who is following up or not. Also, during the discussion, when you teach face-to-face, you can encourage students to participate in the discussion more than online’.* However, three participants reported issues with their library access, as they faced limitations when accessing some references. According to one of them, *’we do not have any issues with teaching tools, but I do not have access to all publishers I need’*.

### Theme 3: Process evaluation - Facilitation and Scheduling

Process evaluation provides information regarding the schedule, method of progress, and education method related to the education program.

SDL sessions are currently taught to a large group of the whole class. The faculty suggested several educational strategies. One participant suggested students’ orientation about SDL sessions. This participant said, *‘Students should receive orientation about what they are supposed to do’.* The participants frequently suggested providing learning resources that are not too long and are written in simple language as stated by one participant, *‘I searched as an instructor an article about this topic that is somewhat easy to understand, not very complicated’.* Two participants recommended a mentoring programme to support students. Small group-based learning was mentioned by some participants as stated by one participant, *‘groups need to be small’*. Delivering SDL in the form of CBL was suggested by one participant. The participant stated, *’Whenever there is an SDL session, we complement it with a sort of CBL’.* The representation of SDL in the curriculum should be increased, according to two participants. One of them stated, *‘we need to increase the number of sessions [SDL]’* Scheduling issues for SDL activities were reported. These issues include rescheduling from the original time or sessions that were scheduled close to summative assessments. One participant explained how scheduling a session close to exam time could negatively impact the student’s preparation for the sessions*: ’[SDL sessions] need to not be close to exam time as [students] will scarify the sessions for the exams’*. In contrast, proper scheduling can be an enabler for SDL. Allowing enough time for students to prepare for the sessions was frequently mentioned by the participants. For example, one participant said *’I used to give them the task to read at the beginning of the week, and SDL is taught the day before the weekend’*.

### Theme 4: Product Evaluation – Students’ engagement and assessment

Product evaluation examines the overall efficacy of an educational intervention and considers the intended and unintended effects. To this end, faculty reflected on whether they felt the SDL sessions met their intended outcomes and the factors that contributed to this. For most of them, the efficacy of the sessions was judged by students’ engagement (or lack of).

It is important to note that all participants were unsatisfied with students’ engagement. One participant noted *’[The students] are not giving it that importance, and that is reflecting in everything, such as their preparations, understandings’.* Similarly, another participant stated: *‘Frankly speaking, students mostly do not like SDL sessions, especially if you are expecting a certain degree of knowledge acquired by them… I would say there are a certain number of students who are very serious, but they are hardly 10% to 15% of the rest of the students’*. Third participant stated, *‘poor engagement; after I give the task to them, no student comes back to me asking about anything’*.

According to three participants, female students tend to engage more. One of these participants stated, *‘I notice girls are very good learners as compared to boys; they are very good learners, and they study hard’* . Two participants mentioned the students who are higher achievers also tend to be more engaged. One of them stated, *‘The more knowledgeable the students are, it is easier for me to deliver the sessions; high GPA students can do very well’* Faculty highlighted a specific gap in the assessment of SDL. In particular, they suggested that the knowledge discussed in SDL sessions is not included in the summative assessment of the internal medicine modules. All participants believed that making marks for SDL topics in the summative exams would improve student engagement and consequently the educational value of the sessions. For example, One participant asserted, *’We need to put marks for attendance and participation in addition to including SDL topics in the exams’*.

## Discussion

The purpose of this study was to investigate the perceptions of faculty regarding SDL activities. The faculty understanding of SDL was in line with Knowles’ definition of SDL in that learners take the initiative and teachers act as facilitators (4). SDL is based on adult learning principles, where adults are problem-oriented and internally motivated (3). Although most participants consider topics that are clinically relevant and common health issues that could address students’ needs and improve intrinsic motivation, which are important elements of SDL (27). However, faculty members were found choosing topics for their sessions rather than problems without students controlling their learning process, and not encouraging peer review or reflection. The participants’ suggestion of faculty training for SDL is consistent with recommendations of recent research. (9, 28, 29). Peer review of teaching is widely used for faculty development and dissemination of best teaching practices in higher education (30).

Regarding learning objectives, several academics have downplayed their significance, claiming that objective-led lessons make students passive and discourage creativity and critical thinking (31, 32); therefore, the educational strategy would be less favourable for SDL. Rather, research on teaching methods indicates that stating learning objectives has a motivational benefit according to Reed (31) and Pelaccia and Viau (33). Furthermore, Knowles (4) recommended that teachers should be able to convert learning needs into precise, attainable, and practical learning objectives to aid learners in setting their own goals as one of the competencies required for SDL.

The findings indicate poor engagement among students, which could be explained by a lack of motivation or skills required for SDL. Previous research supports the latter explanation (i.e. lack of skills), including one study conducted in India (9) and two studies conducted in Saudi Arabia (10, 11). These studies concluded that students desire to learn but need assistance to improve their SDL abilities (e.g. time management skills). This fact emphasises the vital role of educators. In this study, the findings suggest that females have better engagement; consequently, they could be more ready for SDL than males. This finding is in line with two previous studies; one was conducted in three countries (Saudi Arabia, the Philippines, and Thailand) (34), and the other was conducted in Saudi medical schools (10). However, the findings contradict a study conducted in Saudi Arabia that reported no gender discrepancy regarding attitude towards SDL (11). Regarding the academic year, existing studies support the findings in this study, as they revealed that students who had advanced further in their educational programme were more ready for SDL (Slater and Cusick (7), Salih, Sembawa (10).

The faculty suggestions to improve SDL sessions are supported by the literature. Student preparedness was recommended in previous publications (9, 18, 28). SDL is fostered by the involvement of mentors (27). Also, Sheppard-Law, Curtis (35), Aho, Ruparel (36) reported that mentor-guided SDL improves learning. Furthermore, SDL is an active, student-based learning for which small group methods (e.g. TBL) are effective (17, 37). TBL is one of the methods used to implement and enhance SDL (5, 13, 38). In addition, students ascribe more value to educational activities in which theory is linked to real-life practice (33). As a result, CBL enhances students’ motivation towards SDL (39). Hill, Peters (13) reported that case-based TBL is effective in delivering SDL.

Regarding learning environment, The effectiveness of SDL is influenced by a variety of individual and contextual factors (40). According to Hiemstra and Brockett (41), learners are more self-directed when the educational environment is supportive. Today, technology may make learning more accessible and efficient; however, it can also be disruptive when it is poorly designed or used (3). Chakkaravarthy, Ibrahim (8) reported that online learning improves SDLR. In contrast, lack of access to technology can hinder the process of SDL (17). The findings are supported by Douglass and Morris (17), who reported scheduling as a facilitator/barrier for SDL and the importance of proper planning by the administrators. Furthermore, Levett-Jones (6) argued that appropriate arrangements should be made to meet the students’ learning needs and time requirements.

Assessment has a significant educational impact, as students adapt their learning strategies to the assessment methods (42). Therefore, assessment can be used to promote learning (43). This can be achieved through the behaviourist (i.e. summative) approach or the constructive (i.e. formative) approach (3). The participants’ focus was on the behaviourist approach. Summative assessment does not typically support SDL. According to Knowles (4), SDL is inappropriate when learners are externally motivated, in agreement with a study that revealed higher SDLR associated with intrinsic motivation (34). Knowles (4) further explained that teachers should be able to assess learning outcomes in a way that encourages peer evaluation and reflection on learning.

It is important to stress that this study is not without limitations. The success of an evaluative study largely depends on the participation of the relevant stakeholders especially if it is based on the CIPP model that is comprehensive as it considers the complexity of the educational program and allows the involvement of different stakeholders (19). Therefore, the study validity could be improved by data triangulation (i.e. involving the use of different data sources as stakeholders, such as students) (44). The CIPP model could be utilised for both internal and external evaluation as well (19). Hence, the study could be conducted by different investigators/ evaluators such as experts from another department or institute (i.e. investigator triangulation) (44). Also, this study might be strengthened by methodological triangulation (i.e. involving the use of both qualitative and qualitative methods) (44). In addition, member checking, in which participants review the study findings with the researcher, could be considered (23). Another limitation is that the interviews were conducted in English, which was not the participants’ first language. However, qualitative research is more valid when there is as little distance between meanings expressed by participants and meanings in the findings (45).

## Conclusions

This study aimed to evaluate the SDL activities conducted at the Department of Internal Medicine, Faculty of Medicine in Rabigh, King Abdulaziz University, where SDL has been explicitly implemented in the curriculum. Therefore, the findings of the study contribute to the body of existing literature. Most previous research focused on SDLR and did not evaluate the effectiveness of SDL programmes. Qualitative methodology was used to explore the faculty’s perceptions in-depth. The research was based on the CIPP model of evaluation.

In context evaluation, the results indicate that faculty perceived SDL as a reading task that is a pre-requisite for discussions facilitated by the faculty members. The faculty believed that they were poorly prepared to conduct SDL sessions and needed guidance.

The faculty had different perceptions of the objectives of SDL sessions. The main finding was that students should be able to find and utilise the appropriate resources in addition to improving students’ confidence and independence. The faculty believed that SDL sessions aligned with the internal medicine module’s objectives of enhancing knowledge and communication skills with colleagues. Most of the participants believed that SDL activities address the student’s need to become more self-directed learners.

In input evaluation, the participants revealed that their sessions included clinically relevant topics, regardless of whether they had been discussed in previous lectures. The data revealed that participants were satisfied with online platform available for the sessions but some reported issues with direct interaction with students and library access to some resources.

In process evaluation, the faculty suggested the following to improve SDL sessions: students’ orientation, providing learning resources in simple language, students’ mentoring programmes, teaching small groups, CBL, and more SDL sessions. In addition, proper scheduling of SDL sessions is needed for effective delivery of the sessions by allowing enough preparation time for the students and avoiding overlap with exam time.

In product evaluation, poor student engagement in SDL sessions was a major challenge. The faculty believed that students who were female, higher achievers, and more advanced in the academic year were better engaged in the sessions. The results indicate that there was no assessment of the knowledge learned in SDL sessions, and the participants believed that they needed to weigh the content of SDL sessions in the final mark.

Based on the study findings, the following approaches can be implemented to improve the educational value of SDL sessions: faculty preparation, establishing learning objectives, TBL, combining CBL and SDL sessions, assessment for learning rather than of learning and multidisciplinary approach to establish effective learning environment.

Further research should be conducted based on this study such as a study that identifies effective and ineffective facilitation practices for SDL. In addition, the effect of the learning environment on SDL can be studied further. Expanding this CIPP model-based study by including other stakeholders (e.g. students) or evaluators (e.g. external experts) could be considered. A larger scale study should be conducted involving participants from other departments in a school where SDL is delivered differently (e.g. PBL). Finally, a future study should determine the competencies and skills required for students to engage effectively in SDL.

## Data Availability

The datasets generated and analysed during the study are not publicly available due to ethical considerations but are available from the corresponding author upon reasonable request. The interview guide is available upon request from the corresponding author.

